# Prognosis of 1^st^ time revision due to pain following primary total hip arthroplasty - a nationwide cohort study from the Danish Hip Arthroplasty Register

**DOI:** 10.64898/2026.09.09.26361766

**Authors:** Peter Petersen Hald, Saber M. Aljuboori, Søren Overgaard

**Affiliations:** Department of Orthopedic Surgery and Traumatology, Copenhagen University Hospital, Bispebjerg and Frederiksberg, Copenhagen, Denmark; Department of Orthopedic Surgery, Zealand University Hospital, Køge, Denmark

## Abstract

Revision total hip arthroplasty (THA) for persistent or unexplained pain remains controversial. Uncertainty remains regarding the subsequent risk of re-revision following revision THA for pain.

The objectives of this study are to compare the risk of all-cause and cause-specific re-revision following first-time revision THA performed for pain versus aseptic loosening.

This nationwide, register-based cohort study will use data from the Danish Hip Arthroplasty Register. Patients who underwent primary THA followed by first-time revision THA for pain or aseptic loosening between 1995 and 2025 will be included. Patients will be followed from the index revision until re-revision, death, emigration, or end of follow-up. The primary outcome is time to all-cause re-revision. Secondary outcomes are cause-specific re-revision due to pain, aseptic loosening, infection, or dislocation. Cumulative incidence will be estimated using competing-risk methods, and hazard ratios will be estimated using Cox proportional hazards regression.

The study will provide nationwide evidence on the risk of re-revision following revision THA for pain and may inform clinical decision-making and patient counselling.

## Introduction/background

Total hip arthroplasty (THA) is one of the most commonly performed orthopaedic procedures worldwide and is considered the treatment of choice for patients with end-stage hip osteoarthritis (1). Due to demographic changes, increasing life expectancy, and expanding surgical indications to younger and more active patients, the number of procedures has increased and is expected to continue to rise in the coming years (2).

THA is generally associated with high implant survival rates and high levels of patient satisfaction, improving function and quality of life (3). Nevertheless, complications may occur, including infection, recurrent dislocation, or aseptic loosening, which may necessitate revision of the primary THA (4,5). A subset of patients experiences persistent or unexplained pain following surgery and represents a challenging clinical group (6). In some cases, consistent pain may lead to revision surgery despite the absence of clear radiographic findings or identifiable mechanical or biochemical causes (7).

Although previous studies have evaluated patient-reported outcomes, complication rates, and re-revision rates according to the indication for revision THA (8), comparative population-based evidence regarding subsequent implant survival following revision THA for pain remains limited. Improved knowledge of the risk of subsequent re-revision in this patient population is needed to support clinical decision-making, patient counselling, and quality improvement.

## Objective

The primary objective is to compare the risk of all-cause re-revision between patients undergoing revision THA for the indication “pain” and those undergoing revision THA for “aseptic loosening”.

Secondary objectives are to compare cause-specific risks of re-revision, including re-revision due to pain, aseptic loosening, infection, and dislocation, between the two groups.

## Methods

### Study design

The study will be conducted as a nationwide, register-based cohort study. The study will be reported in accordance with the REporting of studies Conducted using Observational Routinely-collected health Data (RECORD) guidelines(9). We will include patients who have undergone a revision THA in both private and public Danish hospitals in the period from January 1^st^ , 1995, to December 31^st^ 2025.

Patients will be followed from the date of the index revision THA until the occurrence of re-revision, death, emigration, or end of follow-up, whichever comes first.

### Data sources

Data for this study is obtained from the Danish Hip Arthroplasty Register (DHR), a nationwide clinical quality database established in 1995 to monitor and improve the outcomes of primary and revision hip arthroplasty in Denmark (10). Reporting to the register is mandatory for all public and private orthopaedic departments performing THA procedures, ensuring full national coverage and high completeness of recorded procedures. In 2024, the DHR had a reporting rate of approximately 98% for primary THA, and 95% for revision THA (11).

The DHR prospectively collects patient-related and surgery-related information, including the patients’ civil registration number (CPR number), indication for primary and revision surgery, date of surgery, laterality, fixation type, surgical approach, implant characteristics, and perioperative complications.

The Danish Civil Registration System is used to obtain information on death and emigration, ensuring virtually complete follow-up of all included patients.

### Study population

The study population consists of patients registered in DHR who underwent a primary total hip arthroplasty followed by a subsequent revision THA performed for the indication of pain (exposed group) or aseptic loosening (reference group) between 1995 and 2025. The index event is defined as the first revision procedure following the primary THA. In 2024, a total of 1,507 revision THAs were performed in Denmark, of which approximately 4% were undertaken with “pain” recorded as the primary indication. During the same period, 396 re-revisions were performed, with 1.5% due to the indication of “pain.” (11) The Danish Hip Arthroplasty Register (DHR) provides the opportunity to investigate outcomes after revision THA in a nationwide, Danish cohort.

#### Inclusion criteria

Patients were eligible for inclusion if they met all of the following criteria:

- First-time revision THA following a primary THA.
- Age ≥18 years at the time of the primary THA.
- Primary THA performed for one of the following indications:
  - Primary osteoarthritis
  - Secondary osteoarthritis
  - Developmental dysplasia of the hip
  - Sequelae of Perthes disease
  - Slipped capital femoral epiphysis
- Index revision performed using a posterior surgical approach.
- Femoral head size of 28 mm, 32 mm, or 36 mm.

#### Exclusion criteria

Patients were excluded if they met one or more of the following criteria:

- Revision THA performed for indications other than pain or aseptic loosening, including:
  - Periprosthetic joint infection
  - Dislocation
  - Periprosthetic fracture
- Missing or unknown indication for revision.
- Missing patient ID
- Missing laterality of surgery.
- Primary THA with a:
  - Metal-on-metal bearing
  - Ceramic-on-ceramic bearing
- Hybrid reverse fixation in the primary THA

### Outcomes

#### Primary outcome

The primary outcome is time to all-cause re-revision, defined as any subsequent surgical procedure involving partial or complete exchange, removal, or addition of one or more prosthetic components, as well as soft tissue procedures following the index revision THA, as recorded in the Danish Hip Arthroplasty Register (DHR). If the first revision was registered as first stage revision of a planned two stage revision, the subsequent revision will not be considered as re-revision.

#### Secondary outcomes

The secondary outcomes include cause-specific re-revision, defined as re-revision due to pain, aseptic loosening, infection, or dislocation, according to the indication recorded in the DHR.

### Statistical analysis

Patients will be stratified into two groups according to the indication for revision surgery. The exposed group will comprise patients who underwent revision due to pain, whereas the reference group will comprise patients who underwent revision due to aseptic loosening. Baseline characteristics will be presented separately for the exposed and reference groups using descriptive statistics.

The cumulative incidence of re-revision at 2, 5, 10, 15, 20, and 25 years will be estimated separately for each group and reported with corresponding two-sided 95% confidence intervals, accounting for death as a competing event using Fine–Gray methods (12). Hazard ratios (HRs) with corresponding 95% confidence intervals will be estimated using Cox proportional hazards regression. The Cox regression models will be adjusted for age, sex, and comorbidity burden as measured by the Charlson Comorbidity Index (CCI), as well as for time of primary THA and fixation technique (13).

The proportional hazards assumption for the Cox regression models will be assessed using visual inspection of log-minus-log survival plots and Schoenfeld residuals. In the event of violations of the proportional hazards assumption, appropriate remedial measures, such as stratified analyses or the inclusion of time-dependent effects, will be considered. In analyses of cause-specific outcomes, death and re-revision due to other indications are treated as competing events.

## Clinical implications

Revision THA performed for the indication of pain remains controversial. This study aims to compare the subsequent risk of re-revision following first-time revision THA performed for pain versus aseptic loosening, in order to provide evidence to support clinical decision-making when considering to complete a revision surgery for this indication.

## Data Availability

All data produced in the present study are available upon reasonable request to the authors

## Ethical considerations

This study is a register-based observational cohort study using prospectively collected data from the Danish Hip Arthroplasty Register. No intervention or contact with patients will take place.

In accordance with Danish legislation, register-based studies that do not involve direct patient contact or biological material do not require approval from a regional research ethics committee. The study will be conducted in compliance with the General Data Protection Regulation (GDPR) and the Danish Data Protection Act.

All data will be handled in a pseudonymized format, and analyses will be performed on secure platforms. Access to data will be restricted to authorized study personnel only. Individual patients will not be identifiable in any publications or presentations resulting from this study. The study will be conducted in accordance with the principles of the Declaration of Helsinki and relevant national guidelines for register-based research. Results will be reported transparently and objectively. Any potential conflict of interest will be disclosed.

## Reporting and trial registration

As a register-based study, this research will be reported in accordance with the REporting of Studies Conducted using Observational Routinely-collected Data (RECORD) statement. The study protocol will be made publicly available.

## Budget

Publication dkr. 25.000

Presentation ISAR congress dkr. 15.000

## Tables

**Table 1.** baseline characteristics.

| Variable | Pain | Aseptic loosening |
| --- | --- | --- |
| THA revision (n) |  |  |
| Age (mean +/- SD) |  |  |
| Gender (% female) |  |  |
| Charlson comorbidity index |  |  |
| Fixation technique |  |  |
| Femoral head size |  |  |
| Date of primary revision surgery |  |  |
| 1995-1999 |  |  |
| 2000-2004 |  |  |
| 2005-2009 |  |  |
| 2010-2014 |  |  |
| 2015-2019 |  |  |
| 2020-2025 |  |  |
| Time from primary THA to first revision (years) |  |  |
| <1 year |  |  |
| 1-5 years |  |  |
| >5 years |  |  |

**Table 2.**
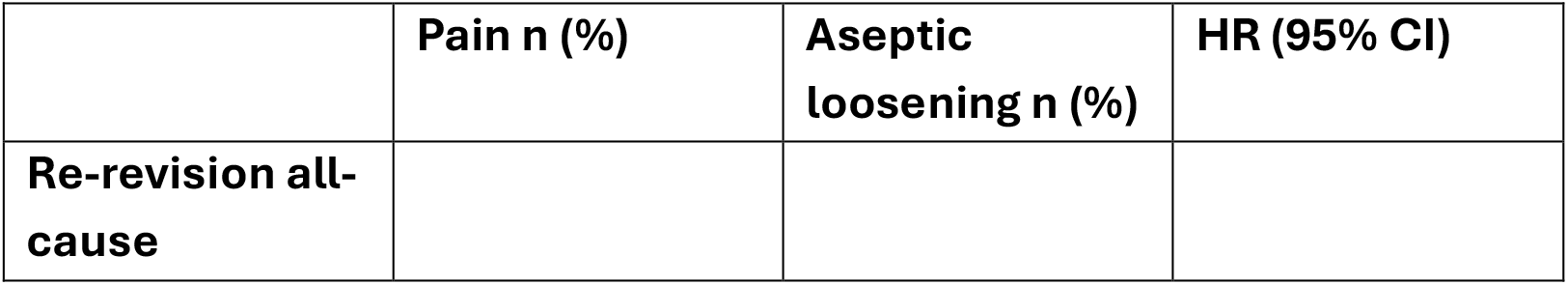
all cause re-revision THA.

**Table 3.**
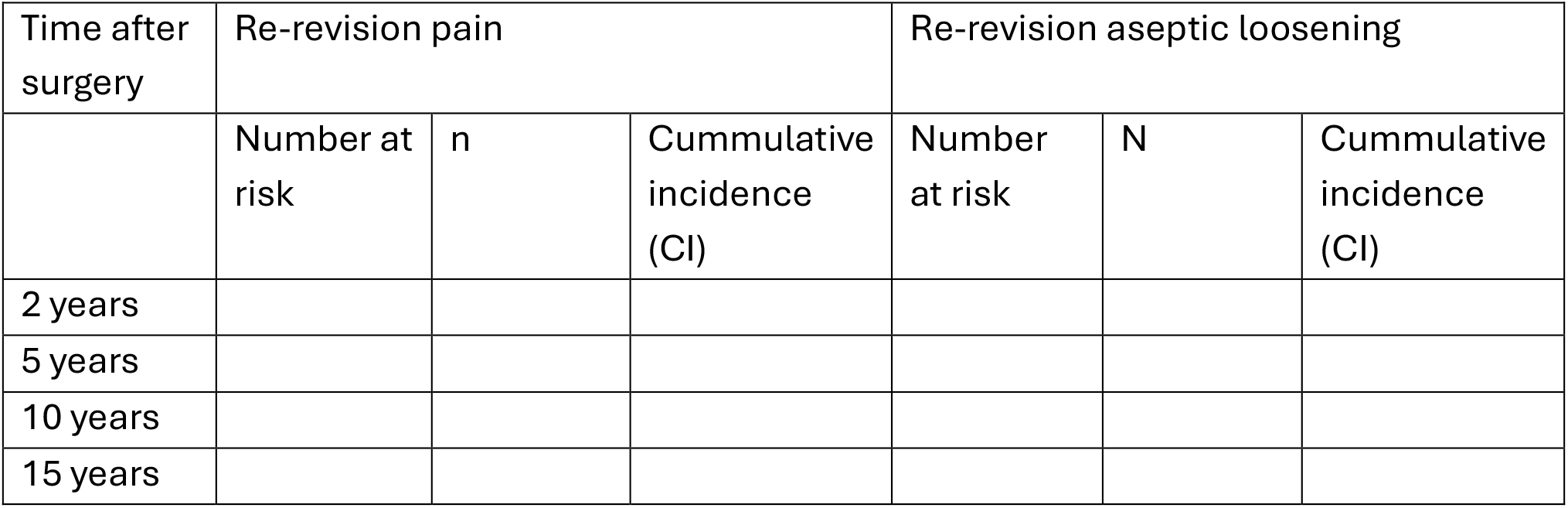
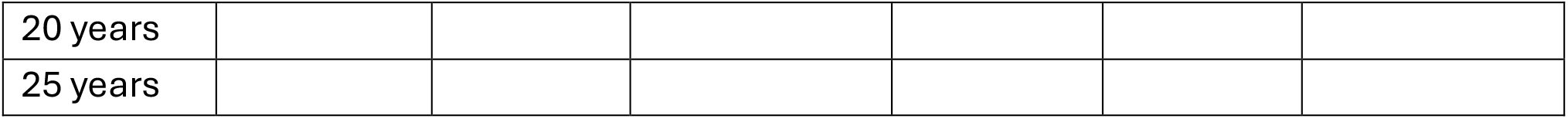
cumulated incidence of THA re-revisions after 2, 5, 10, 15, 20 and 25 years.

**Table 4.** cause specific re-revisions.

| **Indication for re-revision** | **Pain n=** | **Aseptic loosening n=** | **Absolute risk difference (95% CI)** |
| --- | --- | --- | --- |
| **Pain** |  |  |  |
| **Aseptic loosening** |  |  |  |
| **Infection** |  |  |  |
| **Dislocation** |  |  |  |
| **Fracture** |  |  |  |
| **Other** |  |  |  |

## References

1. Hulin K, Fearon A, Newman P. What are the diagnoses attributed to persistent hip pain after hip arthroplasty? A systematic review. J Clin Orthop Trauma. 2025 Aug;67:103036. doi:10.1016/j.jcot.2025.103036

2. Stubnya BG, Schulz M, Váncsa S, Szilágyi GS, Szatmári A, Bejek Z. Global Trends in Joint Arthroplasty: A Systematic Review and Future Projections. J Clin Med. 2025 Nov 19;14(22):8214. doi:10.3390/jcm14228214

3. Laigaard J, Aljuboori SM, Nikolajsen L, Mathiesen O, Lunn TH, Overgaard S. Chronic Postsurgical Pain after Primary Total Hip Arthroplasty for Osteoarthritis: A Nationwide Cross-Sectional Survey Study. J Arthroplasty. 2025 Oct;S0883540325012495. doi:10.1016/j.arth.2025.09.057

4. Oltean-Dan D, Apostu D, Tomoaia G, Kerekes K, Päiusan MG, Bardas CA, et al. Causes of revision after total hip arthroplasty in an orthopedics and traumatology regional center. Med Pharm Rep. 2022 Apr 20;95(2):179–84. doi:10.15386/mpr-2136

5. Cascales V. Why revision of total hip arthroplasty fails: a retrospective consecutive cohort study of 963 patients. Vol. 7. 2026;7(2).

6. Murphy J, Pak S, Shteynman L, Winkeler I, Jin Z, Kaczocha M, et al. Mechanisms and Preventative Strategies for Persistent Pain following Knee and Hip Joint Replacement Surgery: A Narrative Review. Int J Mol Sci. 2024 Apr 26;25(9):4722. doi:10.3390/ijms25094722

7. Classen T, Zaps D, Landgraeber S, Li X, Jäger M. Assessment and management of chronic pain in patients with stable total hip arthroplasty. Int Orthop. 2013 Jan;37(1):1–7. doi:10.1007/s00264-012-1711-6

8. Innocenti M, Smulders K, Willems JH, Goosen JHM, Van Hellemondt G. Patient-reported outcome measures, complication rates, and re-revision rates are not associated with the indication for revision total hip arthroplasty: a prospective evaluation of 647 consecutive patients. Bone Jt J. 2022 Jul 1;104-B(7):859–66. doi:10.1302/0301-620X.104B7.BJJ-2021-1739.R1

9. Benchimol EI, Smeeth L, Guttmann A, Harron K, Moher D, Petersen I, et al. The REporting of studies Conducted using Observational Routinely-collected health Data (RECORD) Statement. PLOS Med. 2015 Oct 6;12(10):e1001885. doi:10.1371/journal.pmed.1001885

10. Gundtoft P, Varnum C, Pedersen AB, Overgaard S. The Danish Hip Arthroplasty Register. Clin Epidemiol. 2016 Oct;Volume 8:509–14. doi:10.2147/CLEP.S99498

11. Madsen KF. Dansk Hoftealloplastik Register. 2024.

12. Van Der Pas S, Nelissen R, Fiocco M. Different competing risks models for different questions may give similar results in arthroplasty registers in the presence of few events: Illustrated with 138,234 hip (124,560 patients) and 139,070 knee (125,213 patients) replacements from the Dutch Arthroplasty Register. Acta Orthop. 2018 Mar 4;89(2):145–51. doi:10.1080/17453674.2018.1427314

13. Charlson ME, Pompei P, Ales KL, Mackenzie CR. A new method of classifying prognostic comorbidity in longitudinal studies: Development and validation. J Chronic Dis. 1987 Jan;40(5):373–83. doi:10.1016/0021-9681(87)90171-8

